# Impact of antibiotics to off-target infant gut microbiota and resistance genes in cohort studies

**DOI:** 10.1101/2021.11.02.21265394

**Authors:** Rebecca M. Lebeaux, Juliette C. Madan, Quang P. Nguyen, Modupe O. Coker, Erika F. Dade, Yuka Moroishi, Thomas J. Palys, Benjamin D. Ross, Melinda M. Pettigrew, Hilary G. Morrison, Margaret R. Karagas, Anne G. Hoen

## Abstract

**Background:** Young children are frequently exposed to antibiotics for otitis media and respiratory infections, with the potential for collateral consequences on the gut microbiome. The impact of antibiotic exposures to off-target microbes (i.e., bacteria not targeted by antibiotic treatment) and antibiotic resistance genes (ARGs) is unknown.

**Methods:** We used metagenomic sequencing data from paired stool samples collected prior to antibiotic exposure and at 1 year from over 200 infants and a difference-in-differences approach to assess the relationship between subsequent exposures and the abundance or compositional diversity of off-target microbes and ARGs while adjusting for covariates.

**Results:** By 1 year, the relative abundance of multiple species and ARGs differed by antibiotic exposure. Compared to infants never exposed to antibiotics, *Bacteroides vulgatus* relative abundance increased by 1.72% (95%CI:0.19,3.24) while *Bacteroides fragilis* decreased by 1.56% (95%CI:-4.32,1.21). *Bifidobacterium* species also exhibited opposing trends suggesting differential antibiotic selection. Overall, antibiotic exposure was associated with a dose-dependent decrease in alpha diversity of off-target microbes. ARGs associated with antibiotic exposure included class A beta-lactamase gene *CfxA6*. Among infants attending day care, *Escherichia coli* and ARG abundance were both positively associated with antibiotic use.

**Conclusion:** Further quantifying impacts to off-target microbes and ARGs has implications for antibiotic stewardship

**Impact:**

- Infants are frequently exposed to antibiotics for respiratory illnesses, but the extent of impact to off-target gut microbiota and antibiotic resistance genes is unknown.
- We quantified these impacts in 2 cohort studies using a difference-in-differences approach. Novel to the microbiome space, this enabled us to use pre/post antibiotic data to emulate a randomized controlled trial.
- Compared to infants unexposed to antibiotics between six weeks and 1 year, the relative abundance of multiple off-target species and antibiotic resistance genes was altered.
- This research highlights the need to consider the microbiome in antibiotic stewardship and offers a new framework for quantifying change.

## Introduction

Infants and young children are exposed to more antibiotics than any other age group^1,2^ with an estimated 30-40% of children receiving at least one course of antibiotics within their first year of life^3–7^. Although most of these antibiotics are prescribed for acute respiratory tract infections including acute otitis media^8,9^, an unintended consequence of antibiotic use is its impact on the abundance and diversity of microbes other than the targeted pathogen^10,11^. Understanding antibiotics’ impacts on off-target or “bystander” microbes in the infant gut, in particular, is significant for two main public health reasons^11^. First, systemic antibiotics can result in microbial dysbiosis as the developing infant gut is sensitive to perturbation^12^. Encompassing both taxonomic and metabolic changes, antibiotics and resulting dysbiosis have been associated with negative health outcomes in young children including overweight/obesity^13– 16^, asthma^7,17^, and celiac disease^18^. Second, antibiotic exposures can lead to antimicrobial resistance. This is a particular concern in the human gut, a known reservoir for antimicrobial resistance genes (ARGs)^19,20^. Indeed, multiple studies have found that antibiotic exposures lead to the proliferation of ARGs that confer resistance to the antibiotic prescribed and to other antibiotics^21–24^.

Mitigation of antimicrobial resistance is a global health priority as the incidence of infections requiring second or third-line broader spectrum antibiotics for treatment is on the rise^25^, leading to further risk of antibiotic resistant microbial infections. Integrated strategies that consider the microbiome are increasingly becoming useful for improving the surveillance of ARGs and providing clinical recommendations to prevent antimicrobial resistant infections^26^. Commensal bacteria can harbor ARGs and carriage prevalence of potentially pathogenic microbes can lead to antimicrobial resistant infections. Thus, there is an urgent need to conduct epidemiologic studies and build quantitative models to understand how antibiotics affect off-target microbes, antimicrobial resistance, and unintended health outcomes.

Few studies have attempted to quantify relationships between antibiotic use, off-target microbes, and the spread of antibiotic resistance^10,27^. These studies were limited in that they were cross-sectional or ecologic in design, did not consider antibiotic exposure and microbiome data from the same individuals longitudinally over time, and did not assess ARGs directly.

The goal of the current study was to quantify the population-level effects of antibiotics on the abundance and diversity of off-target microbes and ARGs. Specifically, we use pre/post antibiotic data to estimate average population-level changes to individual microbes, ARGs, and compositional diversity metrics emulating a randomized controlled trial. This difference-in-differences approach is frequently used in health services and policy research but has not been previously applied to microbiome studies^28^. As antibiotics are often prescribed for respiratory illnesses and ear infections unnecessarily^8,9,29^, this study offers insight that can support antibiotic stewardship practices in light of the effect of antibiotics on commensal gut microbes and ARGs.

## Methods

### Study cohorts

The New Hampshire Birth Cohort Study (NHBCS) is an ongoing prospective cohort study of over 2250 pregnant women and their young children from New Hampshire and Vermont. A detailed description of the cohort has been published^30–32^, but, in brief, enrollment began in 2009 to study the effects of environmental exposures on pregnant women and young children. Extensive data are available for the first year of life and include delivery and pediatric medical records, as well as interview data for 4, 8, and 12 months post-delivery. Stool samples have been collected and undergone shotgun sequencing from a subset of children throughout the first year of life but predominantly at the 6-week and 1-year time points. Pregnant women and their children were recruited using the NHBCS’s current Dartmouth Institutional Review Board approved procedures by the Center for the Protection of Human Subjects.

The DIABIMMUNE Study is a cohort of infants in Finland, Estonia, and Russia focused on the hygiene or microbiota hypothesis and its potential role in the development of autoimmune diseases^33^. We used data from infants in a DIABIMMUNE sub-study that aimed to examine the role antibiotics play in microbiome development over the first 3 years of life^23^. The sub-study included 39 infants from Finland having either 0 or at least 9 antibiotic exposures within their first 3 years of life. Children in this cohort had stool samples collected throughout their early life starting at about 2 months of age. We accessed quality-controlled shotgun sequencing FASTQ files and covariate data through their publicly available website^34^.

Additional information on covariates, the stool microbiome processing pipeline, quality control, and profiling are provided in the **Supplemental Notes** (online).

### Antibiotic exposure classification

#### NHBCS

We were interested in systemic antibiotic exposures between the 6-week and 1-year time points (to match available stool samples) with a goal of studying both exposure (yes/no) and the frequency of antibiotic courses given to all infants. A unique advantage of the NHBCS is that antibiotic prescriptions, including indication, are captured in both medical records and caregiver questionnaires administered at 4, 8 and 12 months of age.

As outlined in **Supplemental Figure S1** (online), not all infants with stool samples available had full medical record or interview data over the first year of life. Therefore, we considered two sub-cohorts for assessing antibiotic exposure. The first cohort, the NHBCS Antibiotic Exposure Cohort, classifies antibiotic exposure from medical record and/or interview data. This cohort was used to assess differences between antibiotic exposure versus non-exposure. In this dataset, we have overall less precise knowledge of the timing of antibiotics (i.e., the exact day of antibiotic prescription) but were able to maximize sample size. Our second cohort, the NHBCS Antibiotic Frequency Cohort, only uses antibiotic prescription data from medical records enabling us to know the number and exact timing of antibiotics prescribed. The schema, sensitivity analysis, assumptions, and sample sizes for these two sub-cohorts are available in the **Supplemental Notes** (online).

#### DIABIMMUNE Study

To increase the external validity of this study and include subjects that were known to have a relatively high number of antibiotic exposures, infants from the DIABIMMUNE Study cohort were included. Antibiotic exposures in the DIABIMMUNE Study were collected through parental reporting of antibiotic use, duration, and reason for use as described previously^33^. Infants from the DIABIMMUNE Study were added to NHBCS sub-cohorts to make combined cohorts, referred to respectively as the Antibiotic Exposure and Antibiotic Frequency Cohorts [**Supplemental Figure S1** (online)].

### Statistical analyses

The difference-in-differences approach is frequently used in health services and policy research to estimate the effect of change due to an intervention or policy on a population when a randomized controlled trial is not possible or ethical^28^. The advantage of using this design is that it uses baseline information (i.e., data from before the intervention) to account for intra-subject variation over time and a comparison group to assess differences among those exposed and unexposed to an intervention. This approach is equivalent to assessing the interaction effect between the exposed group and the post-intervention time point^35^. Assumptions required for the difference-in-differences approach^28,35^ along with modeling frameworks are discussed further in the **Supplemental Notes** (online).

## Results

### Study groups

We used rigorous classification rules [**Supplemental Notes** (online)] to retrospectively assess antibiotic exposure and frequency in 238 infants in the New Hampshire Birth Cohort Study (NHBCS) that had shotgun sequencing data from stool samples collected at both ∼6-week and ∼1-year time points [**Supplemental Figure S1** (online)]. Ultimately, we assigned 183 infants from the NHBCS as exposed or unexposed to antibiotics for a specific condition based on interview and/or medical record data (the NHBCS Antibiotic Exposure Cohort). A group of 99 infants could be classified by antibiotic frequency based on medical record data (NHBCS Antibiotic Frequency Cohort). Sensitivity analyses were used to evaluate exposure misclassification between medical record and interview data, but we found high concordance between the two methods [**Supplemental Notes** (online)]. In addition to infants participating in the NHBCS, we were able to classify the antibiotic exposure and frequency profiles of 33 infants with stool samples collected at approximately 2 months and 1 year from the DIABIMMUNE Study^23^.

### Descriptive characteristics of infants and samples

Comparisons between the DIABIMMUNE and NHBCS Antibiotic Exposure and Antibiotic Frequency sub-cohorts indicated that the proportions of infants that were female and that were born full-term were similar (**Table 1**, *χ*^2^ -*p*-value > 0.1). Infants from the DIABIMMUNE Study were more likely than infants from at least one NHBCS sub-cohort to be vaginally delivered (*χ*^2^ -*p*-value < 0.1). The average baseline age and days breastfeeding of infants from the DIABIMMUNE Study was approximately 20 days greater than that of infants from the NHBCS (Kruskal-Wallis *p*-value < 0.1).

**Table 1:**
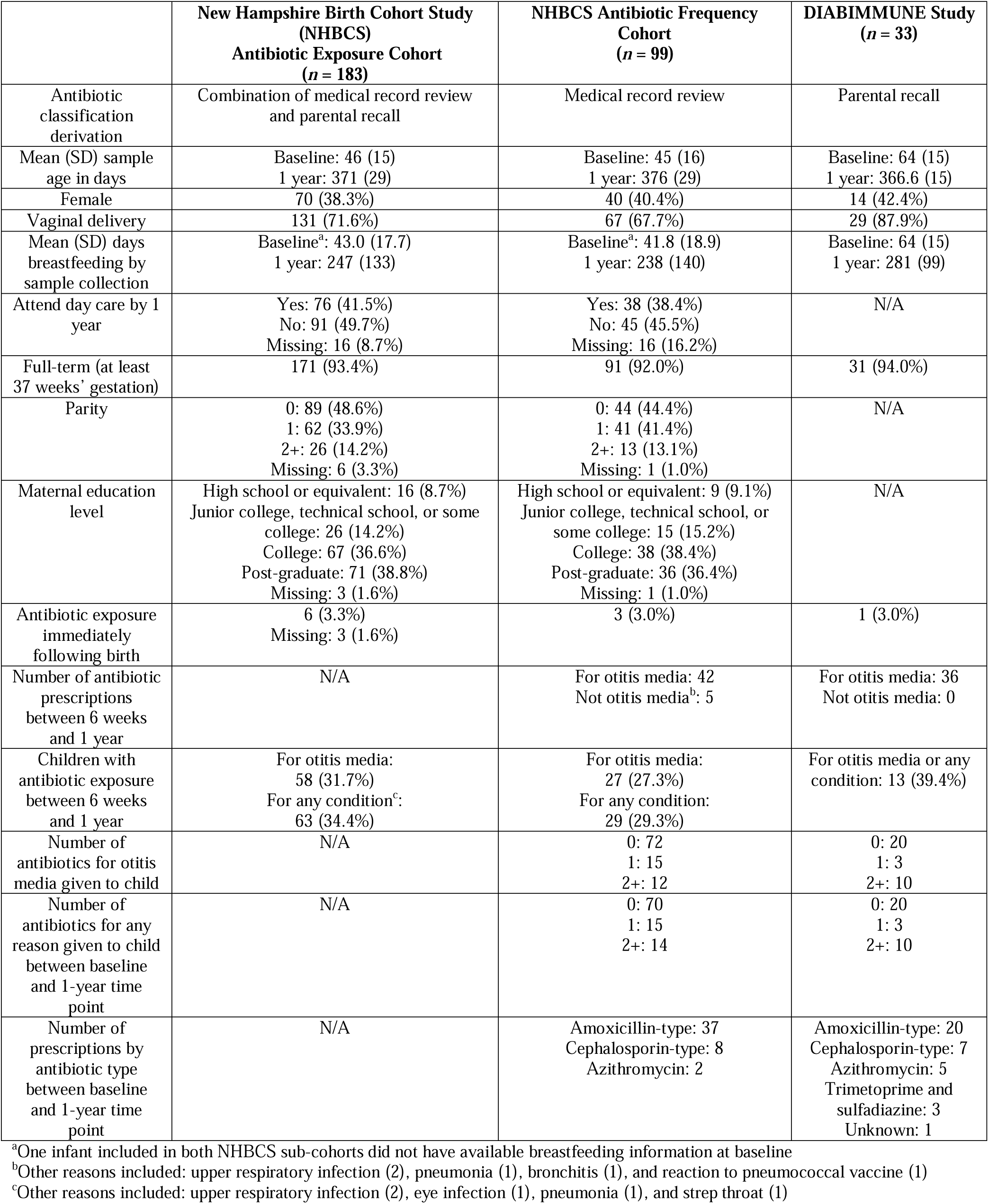
Descriptive overview of infants in the study

The majority of antibiotic exposures occurred after 6 months of age and for otitis media (**Figure 1**). As expected based on the inclusion criteria that DIABIMMUNE Study infants would have high exposure to antibiotics over the first 3 years of life, infants in the DIABIMMUNE Study were more frequently exposed to antibiotics and, when exposed, were given more antibiotics than infants in the NHBCS. Amoxicillin-type antibiotics (e.g., amoxicillin, amoxicillin with clavulanate) were the most commonly prescribed antibiotics among all infants.

**Figure 1:**
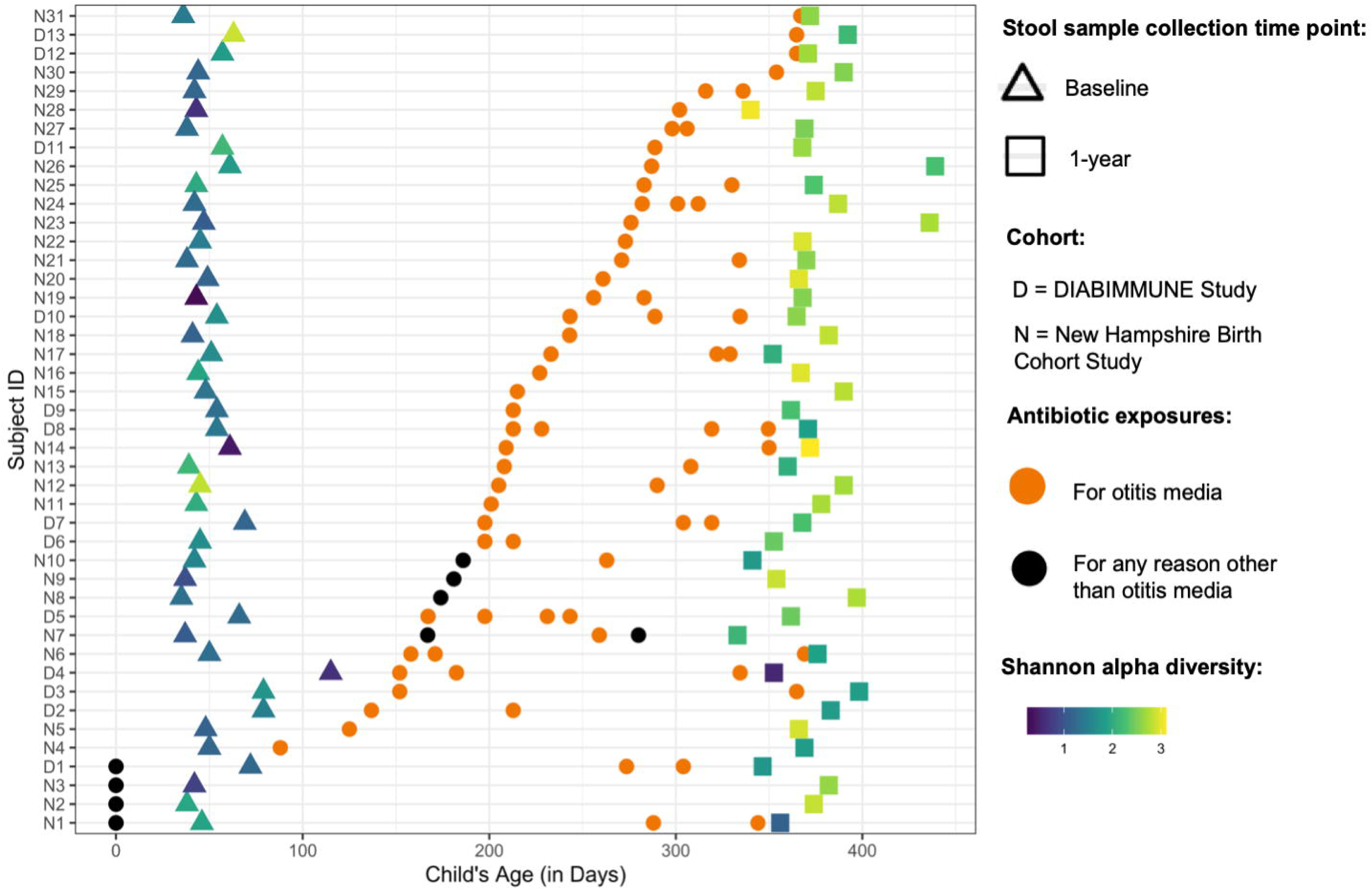
Descriptive overview of antibiotic exposures among New Hampshire Birth Cohort and DIABIMMUNE Study infants exposed to antibiotics in the Antibiotic Frequency Cohort. Figure only includes antibiotic exposures occurring before the 1-year stool sample collection. Samples are ordered by first antibiotic exposure. Baseline samples were collected at approximately 6 weeks for New Hampshire Birth Cohort Study participants and 2 months for DIABIMMUNE Study infants. Age of antibiotic exposure reflects the start day of antibiotic exposure. Infants with antibiotic exposures prior to the baseline microbiome collection received them immediately following birth (age 0 days).

### Abundance of off-target microbes and antibiotic resistance genes

Since all antibiotic exposures were for non-gut associated infections, we classified all microbes as off-target. Looking specifically at antibiotic exposed versus unexposed infants from the NHBCS and the DIABIMMUNE Studies [(**Figure 2A, Supplemental Table S1** (online)], we found the relative abundance change for 2 microbes was significantly different than zero. In particular, the relative abundance of *Bacteroides vulgatus* across the two timepoints increased by 1.72% (95% CI: 0.19, 3.24) more than it would have if the infant was never exposed to an antibiotic, while the relative abundance of *Actinomyces odontolyticus* decreased by 0.008% (95% CI: 0.015, -0.001). Relative abundance changes based on antibiotic exposure went in opposing directions for *Bacteroides* and *Bifidobacterium* species. While *B. vulgatus* abundance increased, *Bacteroides fragilis* decreased by 1.42% (95% CI: -3.06, 0.23). Similarly, *Bifidobacterium bifidum* relative abundance increased 1.41% (95% CI: -0.44, 3.27) more than it would have if the infant was never exposed, but *Bifidobacterium longum* and *Bifidobacterium breve* decreased by 1.56% (95% CI: -4.32, 1.21) and 1.14% (95% CI: -3.55, 1.27) respectively.

**Figure 2:**
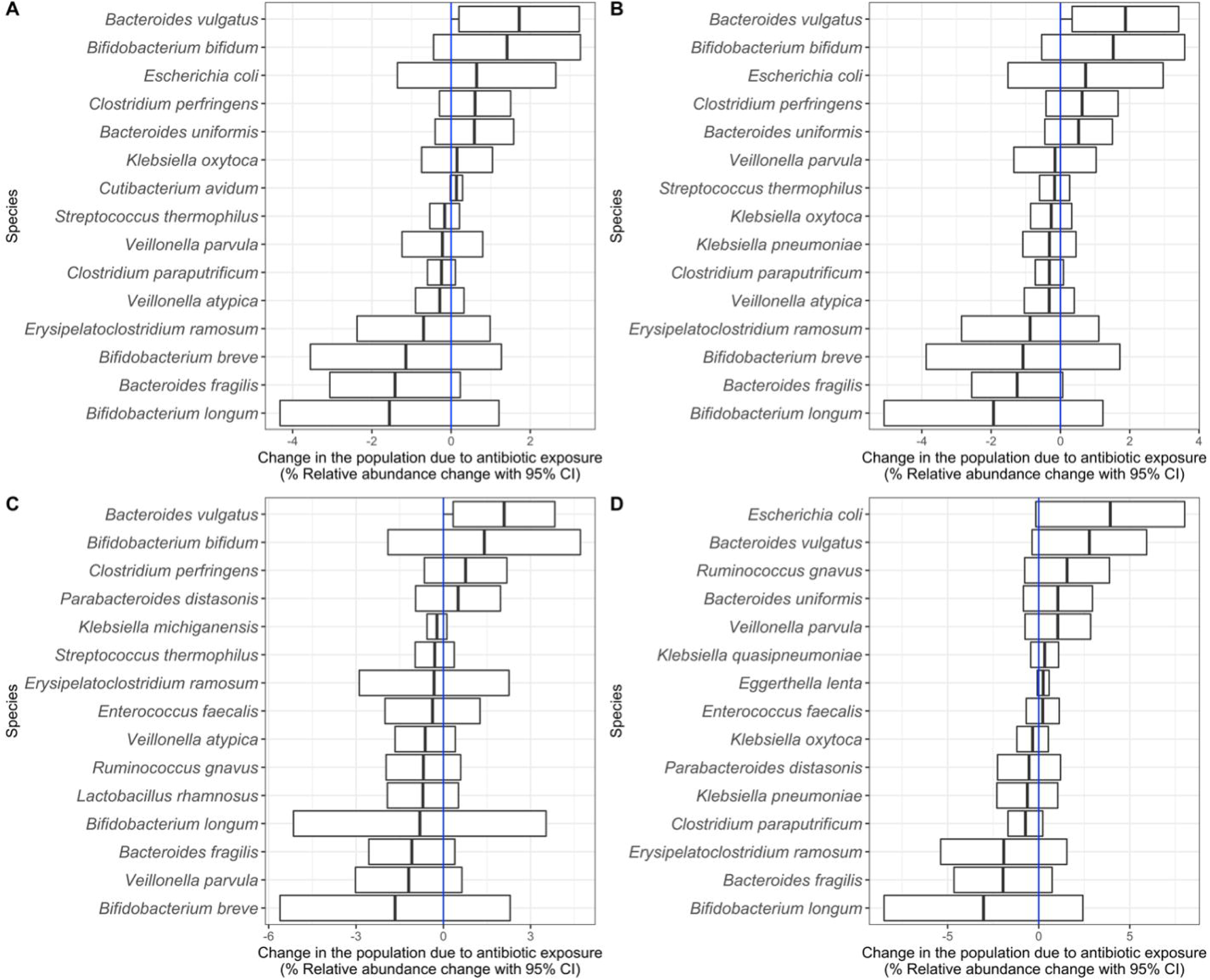
Off-target species changes in the population due to any antibiotic exposure. **A)** Change in the population due to antibiotic exposure in the Antibiotic Exposure Cohort (*n* = 216). The difference-in-difference model for the relative abundance change between exposed and unexposed infants across the two timepoints was adjusted for sample age, duration of breastfeeding, delivery mode, sex, gestational age, antibiotic use immediately following birth, study cohort, and a random effect for each subject. **B**) Change in the population due to antibiotic exposure in NHBCS infants (*n* = 183). **C**) Change in the population due to antibiotic exposure in NHBCS infants that did not attend day care by 1 year (*n* = 91). **D)** Change in the population due to antibiotic exposure in NHBCS infants that did attend day care by 1 year (*n* = 76). **B** was adjusted for the same variables as **A** with the exception of study cohort and addition of day care attendance by 1 year. **C** and **D** models were adjusted for the same variables as **B** except for day care attendance by one year. For all plots, the 15 species with the greatest absolute change are shown. Species are ordered by the point estimate for the relative abundance change.

As day care attendance increases an infant’s exposure to infections^36–38^ and is time variant [see **Supplemental Notes** (online)], we stratified NHBCS infants in the Antibiotic Exposure Cohort by day care attendance by 1 year to attempt to separate the effect of day care attendance and antibiotic exposure on off-target microbes (**Figure 2B-D**). Some microbe associations were consistent in both day care strata (i.e., no evidence of effect modification). However, among infants who attended day care, the relative abundance of *Escherichia coli* and *Veillonella parvula* was considerably higher among infants exposed to antibiotics compared to those who were unexposed, suggesting a synergistic interaction between antibiotic exposure and day care attendance towards the abundance of certain microbes.

We also assessed the impact of antibiotic exposure to changes in ARGs over the follow-up period (**Figure 3**). The most significant change occurred for the Antibiotic Resistance Ontology marker (ARO): 3003097 *CfxA6*, which increased on average by 15.26 reads per kilobase of reference sequence per million samples reads (RPKM) (95% CI: 5.33, 25.20) more than if the infant was not exposed to antibiotics [**Figure 3A**; **Supplemental Table S2** (online)]. Similar results were found among infants only in the NHBCS (**Figure 3B**). Results stratified by day care attendance were discordant for many ARGs. Among infants that did not attend day care by 1 year (**Figure 3C**), *CfxA6* was increased among infants exposed to antibiotics, whereas, a different trend in ARG abundance change was identified among infants that attended day care by 1 year. While *CfxA6* was no longer associated with antibiotic exposure, many other ARGs were positively associated such as *mdtN* (ARO: 3003548), *tetM* (ARO: 3000186), and *mdtO* (ARO: 3003549) (**Figure 3D**).

**Figure 3:**
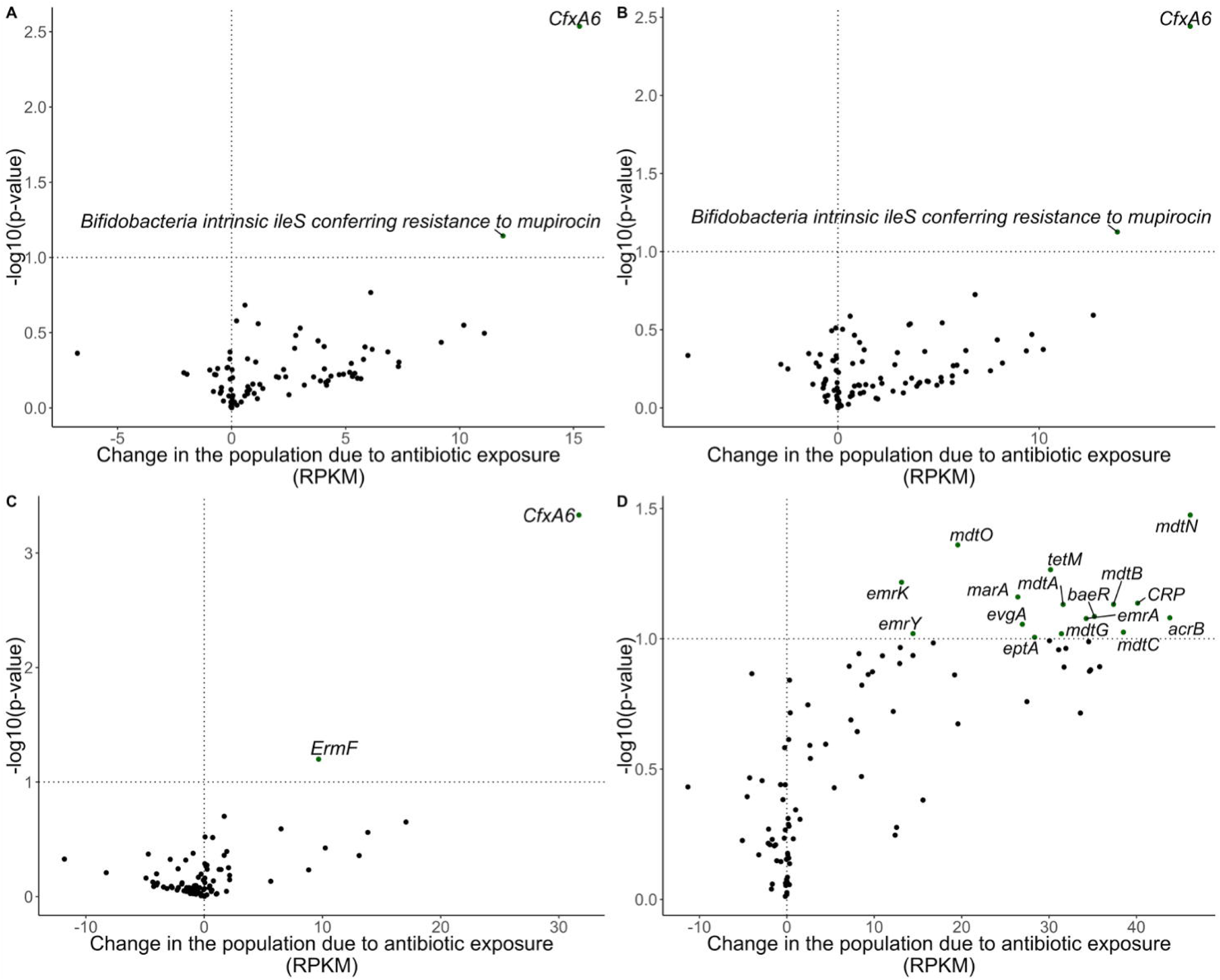
Volcano plots demonstrating abundance of antibiotic resistance gene changes in the population due to any antibiotic exposure in RPKM. Line marks *p*-value < 0.1. **A**) Change in the population due to antibiotic exposure in the Antibiotic Exposure Cohort (*n* = 216). The difference-in-difference model for the relative abundance change between exposed and unexposed infants across the two timepoints was adjusted for sample age, breastfeeding duration, delivery mode, sex, gestational age, antibiotic use immediately following birth, study cohort, and a random effect for each subject. **B**) Change in the population due to antibiotic exposure in NHBCS infants (*n* = 183). **C**) Change in the population due to antibiotic exposure in NHBCS infants that did not attend day care by 1 year (*n* = 91). **D)** Change in the population due to antibiotic exposure in NHBCS infants that did attend day care by 1 year (*n* = 76). **B** was adjusted for the same variables as **A** with the exception of study cohort and addition of day care attendance by 1 year. **C** and **D** models were adjusted for the same variables as **B** except for day care attendance by one year. Only antibiotic resistance genes with a baseline prevalence greater than 20% within each population are included in the plot. RPKM = reads per kilobase of reference sequence per million samples reads

To investigate this further, we looked at overall ARG abundance in the same infants [**Supplemental Table S3** (online)]. In the Antibiotic Exposure Cohort, there was no statistically significant mean ARG difference by antibiotic exposure between time points (516.16 RPKM increase in overall RPKM load among infants exposed to an antibiotic, 95% CI: -904.61, 1936.92) after adjustment for covariates. However, among infants who attended day care in the NHBCS, ARGs increased on average by 2305.54 RPKM (95% CI: -379.80, 4990.87) more in infants exposed to antibiotics than those who were not. For all analyses, while only a few ARGs changed significantly across the two time points, we found that the majority of the estimated mean differences for individual ARGs was greater than 0 suggesting a greater net RPKM trend increase among infants exposed to antibiotics.

To jointly evaluate associations noted in the off-target microbe and ARG analyses, we conducted a block correlation analysis using Hierarchical All-against-All association (HAllA) testing. Across the 41 species and 55 ARGs analyzed, 111 pairs and 47 clusters were identified as having a statistically significant correlation (Benjamini Hochberg *q*-value < 0.05) among unexposed infants and 60 pairs and 19 clusters for exposed infants with 49 pairwise associations overlapping [**Supplemental Table S4** (online)]. Across infants both unexposed and exposed to antibiotics, *E. coli* was highly associated positively and negatively with many ARGs [**Figure 4**; **Supplemental Table S5** (online)]. *Enterococcus faecalis* was statistically significantly associated with multiple ARGs but only among infants exposed to antibiotics.

**Figure 4:**
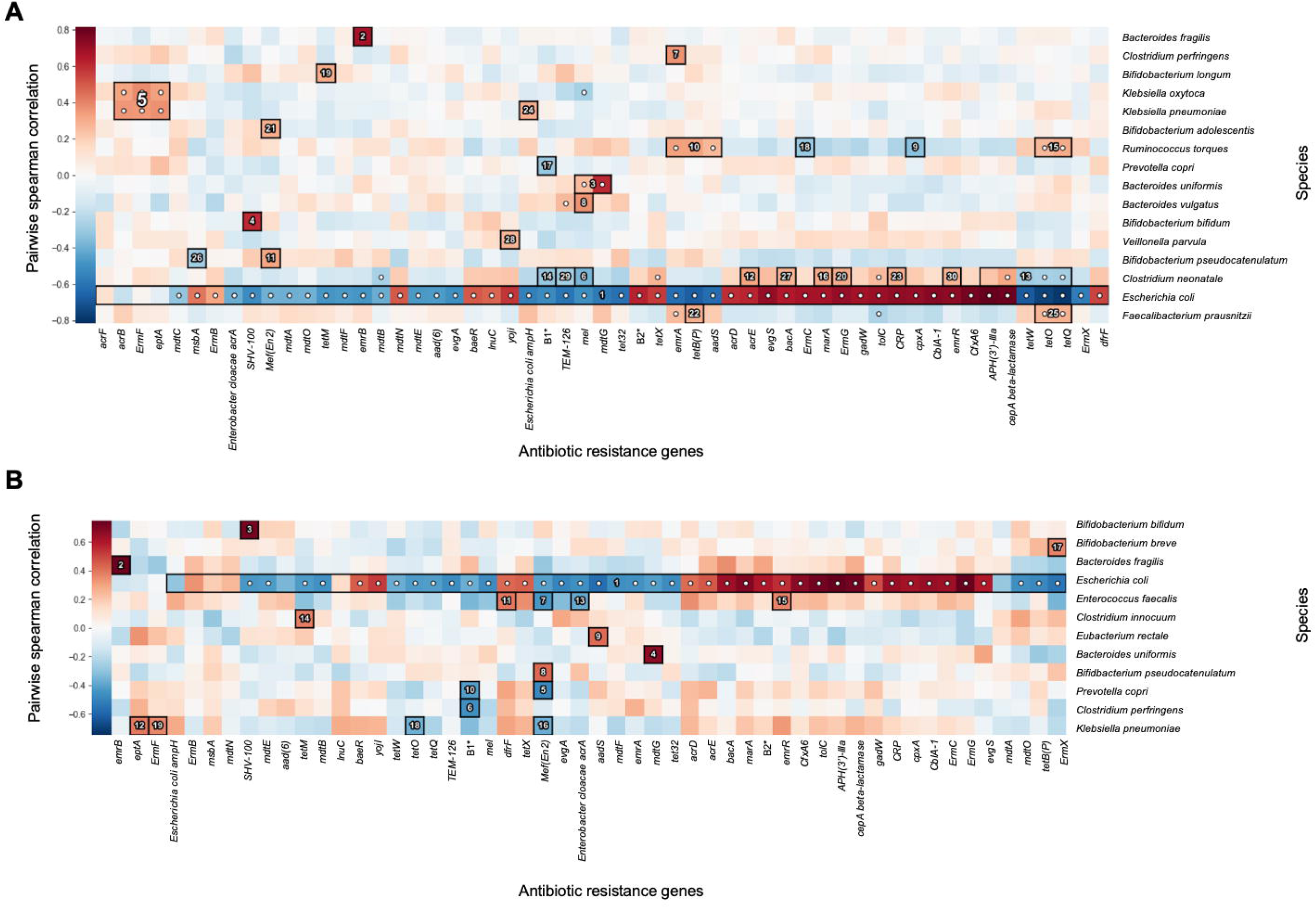
Pairwise spearman correlation between species and antibiotic resistance genes among **A**) infants not exposed (*n* = 140) and **B**) exposed to antibiotics (*n* = 76). HAllA was used to determine associations that were statistically significant. Species and antibiotic resistances were ordered hierarchically. Cluster numbers indicate the rank order of clusters by statistically significant *p*-value. B1*: *Bifidobacterium adolescentis rpoB* conferring resistance to rifampicin, B2*: *Bifidobacteria intrinsic ileS* conferring resistance to mupirocin

### Within (alpha) and between (beta) sample diversity of off target microbes

We evaluated the change to the Shannon index of off-target microbes among infants who had ever received an antibiotic, an antibiotic to treat otitis media, or 2 or more courses of antibiotic between baseline and 1-year measurements. Without accounting for individual random effects, infants in the Antibiotic Exposure Cohort (*n* = 216), had on average a 0.24 (95% CI: - 0.44, -0.05) decrease in alpha diversity more than they would have had they not been exposed across the two time points (**Figure 5A**). This estimate was consistent when random intercepts were incorporated in adjusted linear mixed-effects models that considered any antibiotic exposure or an antibiotic exposure specifically for otitis media [**Supplemental Table S6** (online)]. Likewise, the trend was consistent in analyses restricted to NHBCS infants and subsequently stratified by day care attendance at 1 year [**Supplemental Table S7** (online)]. We also detected a significant dose-dependent association; infants exposed to 2 or more antibiotics between baseline and one-year measurements had on average a decrease in Shannon alpha diversity of 0.34 (95% CI: -0.63, -0.04) more than they would have had they not been exposed.

**Figure 5:**
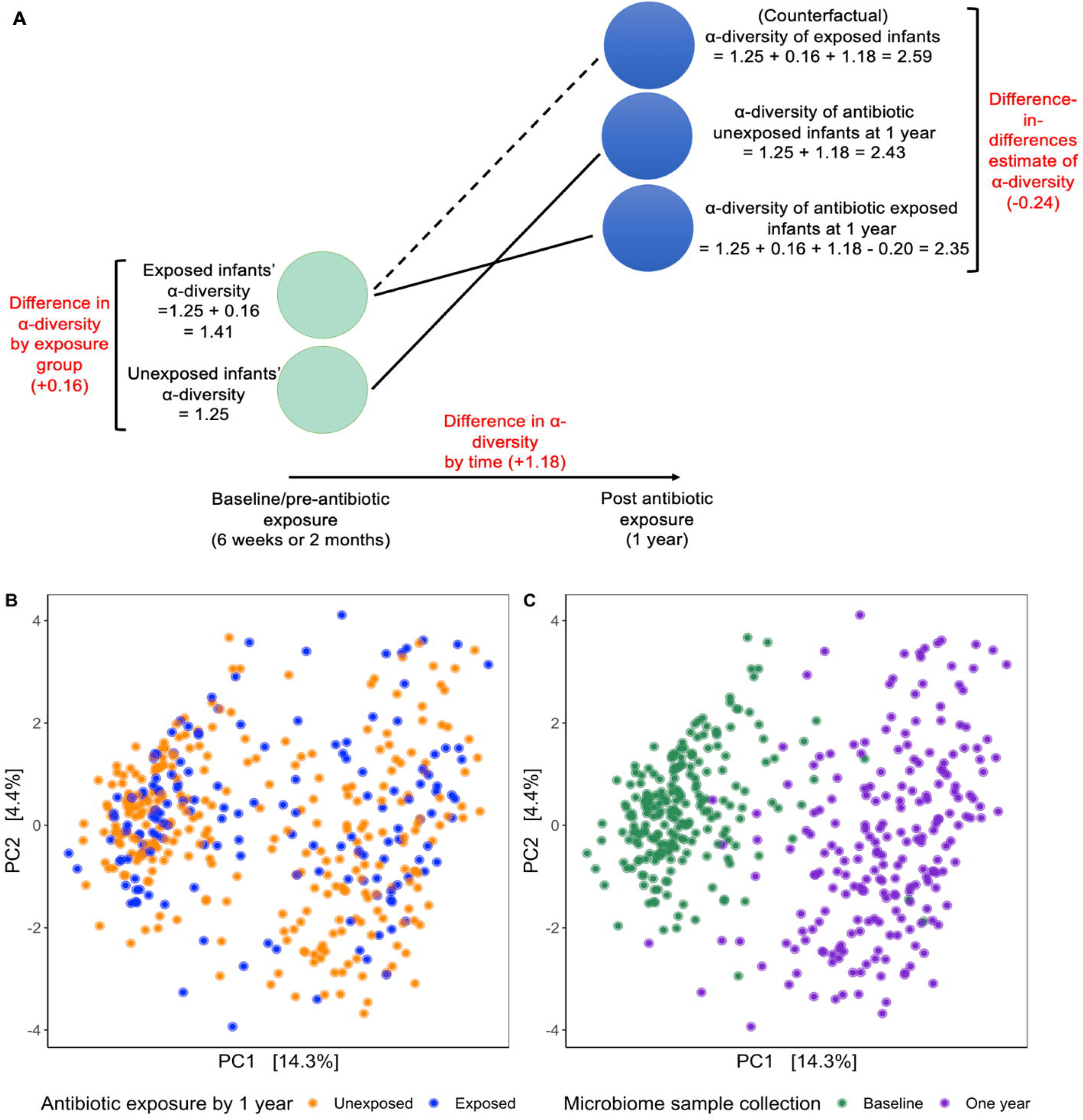
Within and between sample species diversity by antibiotic exposure and time period in 216 infants. **A**) Average within sample (alpha diversity) difference by antibiotic exposure (yes/no) and time period (pre/post antibiotic exposure). Estimates derived from a crude linear regression model with the Shannon index per sample (*n*= 432) as the outcome and antibiotic exposure ever, baseline/1-year treatment, and the difference-in-difference estimate for the interaction as independent variables. All differences are statistically significant at α = 0.05. **B** and **C**) Principal component analysis of between sample diversity. The relative abundance of species was centered log-transformed and plotted using the Euclidean distance. Samples are colored by **B** antibiotic exposure by 1 year (excluding antibiotic exposure immediately following birth) or **C** age of the infant at sample collection.

Results from between sample (beta) diversity analyses of microbiome composition did not visually reveal any clear separation based on antibiotic exposure (**Figure 5B)**, but as predicted, showed the strong impact of sample age on between sample dispersion (**Figure 5C**). While visual results did not depict a strong impact of antibiotic exposures, variation based on the interaction between antibiotic exposure and age were consistently significant across models assessing any antibiotic exposure [**Supplemental Table S8** (online), *p*-value < 0.01)]. Among 216 infants, the interaction between sample age and antibiotic exposure by 1 year described 2.6% of the variation. However, the interaction for exposure to 2 or more antibiotics and age among 132 infants was neither strong nor statistically significant (R^2^ = 0.003, *p*-value = 0.22).

## Discussion

We evaluated the off-target effects of antibiotic exposures on the infant gut microbiome and resistome among a large number of infants from the United States and Finland. Using metagenomic sequencing of pre/post antibiotic stool samples combined with detailed covariate information from 2 separate cohorts, we identified and quantified the population-level impact antibiotics have on off-target microbes and ARGs within the infant gut. We found that even with a broad categorization of antibiotic exposures between approximately 6 weeks and 1 year of life, antibiotics were associated with changes to off-target microbe and ARG abundance.

In the NHBCS Antibiotic Exposure Cohort, we found that 34.4% of infants were exposed to antibiotics between approximately 6 weeks and 1 year of life with 58 out of the 63 (92.1%) antibiotic exposures prescribed for otitis media. This largely correlates with other studies from similar demographic groups that have estimated approximately 30-40% of infants are exposed to antibiotics in the first year^3–7^. Current protocols in the United States recommend antibiotics for most otitis media and lower respiratory tract infections but not for upper respiratory infections^39,40^. Due to challenges in diagnosis, gastrointestinal consequences of antibiotics, and concern for the growing threat of antimicrobial resistance, many country-level antibiotic stewardship protocols encourage watchful waiting^41^. Accordingly, randomized controlled trials^29,42^ and population-level cross-sectional studies^27,43^ have examined delayed, shortened, or no antibiotic therapy for uncomplicated otitis media and respiratory infections^29,42^. While these studies are critical, cohort studies with combined covariate and microbiome data also yield valuable evidence in support of antibiotic stewardship practices because they reflect current treatment practices. Thus, while this study’s main goal was to provide a population-level overview of the impact of antibiotics to infant gut microbiota and the resistome, it also provides public health professionals and clinicians with quantifiable results that may be applicable to antibiotic stewardship recommendations.

Studies inconsistently identify particular commensal microbial abundances that have increased or decreased as a result of antibiotic exposure^44^. While the majority of our results did not indicate statistically significant differences between antibiotic exposed and unexposed infants, a few findings stand out. First, *Bifidobacterium* species exhibited conflicting trends in our study. The relative abundance of two species, *B. longum* and *B. breve*, consistently decreased in the antibiotic exposed population. However, *B. bifidum* increased in relative abundance after antibiotic exposure. This trend suggests that antibiotic exposure may lead to long term selection changes to favor certain species of *Bifidobacterium*. Second, we consistently identified that *B. vulgatus* relative abundance increased in antibiotic exposed children but *B. fragilis* decreased. These results are interesting in context to the DIABIMMUNE Study that identified *B. fragilis* as a “single-colonization species” and *B. vulgatus* as a “multiple-colonization species” based on the dominance of one vs. multiple strains across time^23^. While we did not assess strain diversity, our results suggest that *B. fragilis* may be more susceptible to antibiotics than strains from *B. vulgatus* possibly due to differences in strain-level diversity. An alternative hypothesis could be variation in ARG carriage by species. The Comprehensive Antibiotic Resistance Database (CARD) continually tracks the prevalence of antibiotic resistance phenotypic profiles in reference sequences for many non-pathogenic microbes. According to the CARD prevalence 3.0.9 update, there is wide variability in ARG phenotypes between commensals from the same and different genera^45^. These data, in combination with the results from this study and others assessing antimicrobial susceptibility in bacteria often considered off-target^46,47^, provide further evidence that intra- and inter-species variation is important to consider.

Among infants who attended day care by 1 year, we found that *E. coli* relative abundance increased in antibiotic exposed infants suggesting a synergistic interaction between antibiotic exposure and day care attendance. Although we were unable to identify other studies that assessed both antibiotic exposure and day care attendance, given *E. coli*’s role in horizontal gene transfer^48,49^ and previous studies in the NHBCS and others noting their association with harboring antibiotic resistance genes^21,50–53^, the corresponding changes to resistome composition aligned with changes to *E. coli* relative abundance.

Multiple studies have assessed how antibiotic exposure after the neonatal period impact the resistome^21–23,52,54–57^ but have been conducted in predominantly preterm^21,22^ infants or contained relatively small cohorts (<30 infants) of antibiotic exposed infants^23,54–57^. Interestingly, two studies^21,23^ also identified *CfxA6* beta-lactamase gene and one study^52^ identified *CfxA2* (another *CfxA* class A beta-lactamase gene^58^) as ARG markers of interest. *CfxA* genes have been identified in *Bacteroides, Prevotella*, and *Capnocytophaga* with point mutations differing between *CfxA2* and *CfxA6*^58–60^. The DIABIMMUNE Study found the *CfxA6* gene marker persisted in children long after antibiotic exposure^23^. Although some of these infants were included in our study, we independently found that the gene increased in abundance in infants in the NHBCS only (**Figure 3B**). The most comparable study to ours demonstrated in 662 Danish children that antibiotic exposures during the first year of life impacted the infant gut resistome of healthy infants^52^. In particular, they found time since antibiotic exposure and number of antibiotics influential to overall ARG abundance. Additionally, they identified specific ARGs including *CfxA2* and *APH(3’)-llla* to be increased among infants exposed to a beta lactam inhibitor compared to infants not exposed to any antibiotic. For *CfxA2*, they found that it was also increased among infants given another antibiotic compared to no antibiotic exposure. Although they did not have resistome measurements taken before antibiotic exposures to establish baseline ARG abundances, the results of our study aligned with these results and those presented previously in our cohort^53^.

Previous studies have found mixed results associating diversity and antibiotic exposure depending on geography, age of the child, type of antibiotic prescribed, and time since antibiotic exposure, but, generally, antibiotic exposures are associated with stable or decreased within-sample diversity^44^. Consistently, we found NHBCS infants had a statistically significantly (*p*-value < 0.05) higher alpha diversity than samples in the DIABIMMUNE Study. This may be due to true differences in alpha diversity between the cohorts or lower sequencing depths for samples in the DIABIMMUNE Study. In the DIABIMMUNE cohort study^23^, on average, infants without antibiotic exposures had a microbiome with higher richness, but this was only evident after the first year of life. In our study, we consistently found that there was an inverse association between antibiotic exposure and Shannon alpha diversity prior to 1 year. Interestingly, however, at the baseline timepoint (at approximately 6 weeks of life) infants who were ultimately exposed to antibiotics had higher alpha diversity than infants not exposed to antibiotics. This emphasizes the importance of considering timing of exposure, in particular pre- and post-antibiotic exposure measurement, to discern trends over time.

While this study is one of the largest to consider the population-level impact of antibiotic exposures to the infant gut over a follow-up period, we note some important limitations. Our main limitation was the precise timing and dosage of antibiotic exposures in the NHBCS Antibiotic Exposure Cohort as well as limitations to generalizability among participants in New Hampshire and Vermont^53^. To mitigate this, we utilized stool samples from the DIABIMMUNE Study as a validation cohort since it obtained both precise timing and dose of antibiotic exposures [**Supplemental Notes** (online)]. We also used exposure data with more precision on a smaller sample group of infants via a medical record review to assess timing and type of antibiotic exposure. Although statistical tools have used mixed-effects models to assess microbiome composition longitudinally^61,62^, the difference-in-differences approach offers a straight-forward interpretation of the effect of an intervention in a population through its strong counterfactual estimate of the effect of eliminating unnecessary antibiotic exposures on the infant gut microbiome and resistome. Moreover, the epidemiologic framework provides the flexibility for researchers to estimate the difference-in-differences using a variety of transformations, statistical packages, and tools. A major consideration, however, in formally applying the difference-in-difference approach to microbiome analyses is fully addressing inherent difference-in-difference assumptions^28,35^. For instance, in this analysis we were unable to fully account for the common trend assumption due to only having 2 timepoints to assess metagenomic data. However, in the process of validating our assumptions, sensitivity analysis revealed that antibiotic exposure did differentially impact some microbes and ARGs in infants depending on their day care status; a novel finding. Lastly, an important caveat to much microbiome data are inherent limitations in accurately reflecting the composition of gut microbiota and ARGs using stool samples. Thus, while we did identify the relative abundance of microbes and ARGs changing in our differential abundance analyses, we cannot rule out that the change would affect the absolute abundance, rule out unmeasured confounding, nor fully predict changes in function or the gut itself^63^. Regardless, misclassification likely would be non-differential by antibiotic exposure and we still can conclude that the infant gut microbiome of antibiotic exposed and unexposed infants is different.

Our results support the notion that microbiota and the resistome should be considered when weighing the costs and benefits of antibiotic interventions. Moreover, we apply an established framework used to emulate a randomized controlled trial to the microbiome space. This enabled quantification of the magnitude of the population-level effects of antibiotic exposure in the context of other pressures impacting the developing microbiome in early life. We imagine that this novel application can be used to evaluate the potential impact of new or current antibiotic prescription practices on gut microbiota and residual impacts to ARGs.

## Supporting information

STROBE Checklist

## Data Availability

NHBCS whole metagenomic shotgun sequencing samples are available through the National Center for Biotechnology Information (NCBI) Sequence Read Archive: https://www.ncbi.nlm.nih.gov/sra (Accession number: PRJNA296814). Analytic R markdown scripts are available upon reasonable request.

## Acknowledgements

We have the upmost gratitude for participants, family members, coordinators, clinicians, and researchers involved in the New Hampshire Birth Cohort. We thank Dr. Scot Zens, Terran Campbell, Dr. Weston Viles, and Dr. Jie Zhou for their thoughtful comments and contributions. We would also like to thank the DIABIMMUNE Study for facilitating open-source data sharing.

