## Supplemental Notes and Figure S1 for "Impact of antibiotics to off-target infant gut microbiota and resistance genes in cohort studies"

**Table of Contents**

***Supplemental Figure:***

**Figure S1:** Description of study inclusion criteria and schema for infants………………..……...2

***Supplemental Text:***

**Antibiotic exposure classification and sensitivity analyses – methods**……………………......3

**Antibiotic exposure classification and sensitivity analyses – results**……………………….....7

**Availability of data and materials**……………………………………………………………..11

**Covariates**……………………………………………………………………………………….11

**Stool microbiome assessment**…………………………………………………………………..12

**Statistical analyses**……………………………………………………………………………...14

**Difference-in-differences assumptions and sensitivity analyses**………………..……………18

**References**………………………………………………………………………………….........22

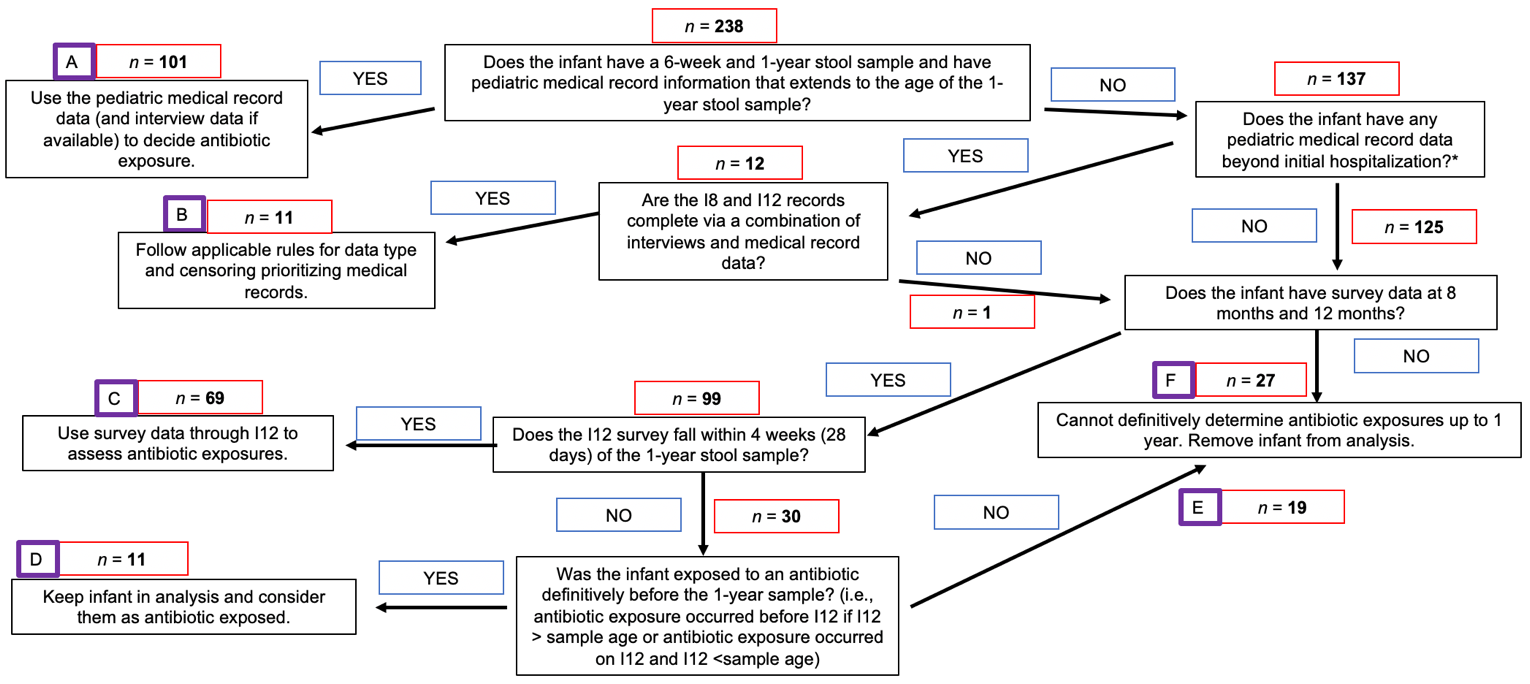

**B**

**C**

**A**

**Figure S1:** Description of study inclusion criteria and schema for infants. **A**) Flow diagram depicting how infants could be included based on antibiotic exposure categories. *Medical records were available during the immediate post-delivery period for all infants. **B**) Depiction of sample size and additional exclusion criteria in both the New Hampshire Birth Cohort and DIABIMMUNE studies. **C**) Combined cohort sample sizes. I8 = 8-month interview, I12 = 12-month interview.

**Antibiotic exposure classification and sensitivity analyses – methods**

Infants in the NHBCS have a range of questionnaire and pediatric medical record data available over the course of their first year of life. Pediatric medical record data is derived from medical record review from a trained professional and includes detailed antibiotic information on the timing, type, and reason for antibiotic prescriptions. Antibiotic exposure is assessed on interviews occurring at approximately 4, 8, and 12 months of life and asks caregivers to identify the type and reason that an antibiotic was given to a child. Through assessment of infants within the NHBCS, we identified that infants with paired 6-week/1-year stool samples had various completion levels for both pediatric medical record and interview data through the first year of life. We classified antibiotic exposure in two ways in an attempt to maximize sample size and minimize exposure misclassification. Thus, we created one cohort that would be used to assess antibiotic exposure vs. no exposure (NHBCS Antibiotic Exposure Cohort) and the other that would assess type and number of antibiotic prescriptions (NHBCS Antibiotic Frequency Cohort). Sample sizes including a flow diagram (**Figure S1**) and results from sensitivity analyses are available below.

*NHBCS Antibiotic Exposure Cohort:*

The conceptual exposure for this cohort was antibiotics given to infants between the baseline (6-week) and 1-year stool sample collection that could impact the gut (i.e., ingested or administered systemically).

Operationally, the antibiotic exposures cohort required more protocols to ensure that antibiotic exposures classified via pediatric medical records and interview data were captured correctly. Depending upon the data available, these were the protocols used to assign infants’ antibiotic exposures:

- If the infant had both pediatric medical record and interview data available, only include infants that had concordant pediatric medical record and interview data indicating an antibiotic exposure.
- If the infant had only interview or pediatric medical record data but that indicated an antibiotic exposure during the time period of interest, mark them as antibiotic exposed.
- If the infant had only interview or pediatric medical record data but that indicated no antibiotic exposure during the time period of interest, mark them as antibiotic unexposed.

Determining if the antibiotic exposure occurred before or after the stool sample was also considered. With pediatric medical record data, the time (days since birth) of the antibiotic prescription was given providing antibiotic exposure timing. However, antibiotic exposures as measured on the interview at 12 months could occur slightly before or after the stool sample was collected. Based on these challenges, we created a schema to determine how to include infants based only on interview data or pediatric medical record data. Since prescription dates are given for pediatric medical record data, the time of antibiotic prescriptions are pulled from there. With interview data, one of the following conditions needed to be met in order to include an infant:

1) 12-month interview data collected within a set time interval before or after the stool sample was collected.
2) 12-month interview occurred significantly earlier than the 1-year stool sample but there was definitive evidence of an antibiotic exposure prior to the stool sample collection.
3) The 12-month interview occurred significantly after the stool sample was collected but no antibiotic exposures were noted.

The next step was determining the time interval to determine correct classification of samples. To determine the best time interval, we utilized infants that had 8- and 12-month interview data as well as pediatric medical record data beyond their first year of life as a sensitivity cohort. We assessed a 14, 21, and 28-day interval around the stool sample collection time point noting concordance and Kappa.

Finally, we applied the chosen time interval and conditions to the full dataset of infants. Based on the sensitivity analysis, the following assumptions were made:

1) 28 days (inclusive) on either side of the stool sample collection point was considered the buffer of inclusion for full records for interview data.
2) The pediatric medical record is considered the “gold standard” because it includes more detailed information on the exact date of the antibiotic prescription which cannot be assumed by the interview. Additionally, parents are less likely to be as knowledgeable about specific antibiotic prescriptions given compared to clinicians.
3) Infants prescribed an antibiotic on the pediatric medical record were actually given an antibiotic.
4) If interview data mentions an antibiotic was given but it’s not matched on the pediatric medical record, it was considered an antibiotic exposure, but ultimately that infant would be censored as there is disagreement between the medical record and interview (i.e., the assumption is that no antibiotic prescriptions on the medical records were missed among infants with full medical records).
5) Infants noted as receiving an antibiotic on the interview data, were given the antibiotic.
6) On the interview, we assume recall bias is minimal as parents would likely know if an infant is given an antibiotic for a condition or not (i.e., there’s a high likelihood that parents would remember either way). There was concern about recall bias for specificity of conditions. In other words, on a medical record the clinician would often say an antibiotic was for “otitis media” while on the interview it may list a more general list of symptoms for why the antibiotic was given (e.g., ear infection, runny nose, and fever). While this doesn’t change the fact that the infant was exposed to an antibiotic, it does limit the ability to compare between the two classification systems.
7) Infants only needed a completed interview at 8 and 12 months as the vast majority of antibiotic exposures occurred after approximately 6 months.
8) Infants that had an antibiotic exposure noted on the 4-month interview were exposed to an antibiotic after the baseline sample was collected. This assumption was based on the sensitivity cohort in which no infant with pediatric medical record data had an antibiotic exposure before 6 weeks unless they were initially given one at birth which is noted in the infant medical record.

*NHBCS Antibiotic Frequency Cohort:*

The conceptual exposure in this cohort was based on the number of antibiotic prescriptions. This was operationalized by assessing all antibiotic prescription from all infants with pediatric medical records up to the 1-year stool sample collection.

Ultimately, the decision was made to include infants that have a recorded antibiotic prescription even if it was not noted on the interview data. We made this assumption because we do not know in infants with missing interview data if 1) the antibiotic prescription would have been captured, 2) antibiotic prescriptions are missing from interview data due to the infant not redeeming their antibiotic prescription, or 3) the caregiver did not recall an antibiotic exposure for his/her/their child.

**Antibiotic exposure classification and sensitivity analyses – results**

*NHBCS Antibiotic Exposure Cohort:*

To assess the time interval that could be used to count antibiotic exposures among infants with interview data in NHBCS Antibiotic Exposure Cohort, we assessed concordance between interview and medical record data at 14, 21, and 28 day intervals among infants with both interview and pediatric medical record data available. 81 infants had pediatric medical record data at least 28 days beyond when their 1-year stool sample was collected as well as having an 8-month and 12-month interview available.

|  |  | **Pediatric medical record** | |
| --- | --- | --- | --- |
|  |  | **Exposed to antibiotic** | **Unexposed to antibiotic** |
| **Interview occurred within 28 days of stool sample or infant could be classified based on other condition*** | **Exposed to antibiotic** | 22 | 1 |
|  | **Unexposed to antibiotic** | 2 | 43 |

Concordance: 65/68 = 95.6%, Kappa = 90.2%

|  |  | **Pediatric medical record** | |
| --- | --- | --- | --- |
|  |  | **Exposed to antibiotic** | **Unexposed to antibiotic** |
| **Interview occurred within 21 days of stool sample or infant could be classified based on other condition*** | **Exposed to antibiotic** | 22 | 0 |
|  | **Unexposed to antibiotic** | 2 | 32 |

Concordance: 54/56 = 96.4%, Kappa = 92.6%

|  |  | **Pediatric medical record** | |
| --- | --- | --- | --- |
|  |  | **Exposed to antibiotic** | **Unexposed to antibiotic** |
| **Interview occurred within 14 days of stool sample or infant could be classified based on other condition*** | **Exposed to antibiotic** | 22 | 0 |
|  | **Unexposed to antibiotic** | 2 | 28 |

Concordance: 50/52 = 96.2%, Kappa = 92.2%

*Other conditions included: 12-month interview occurred significantly earlier than the 1-year stool sample but there was definitive evidence of an antibiotic exposure prior to the stool sample collection. The 12-month interview occurred significantly after the stool sample was collected but no antibiotic exposures were noted.

Using the sensitivity cohort for the 28-day interval (i.e., the cohort with full pediatric medical record and interview data above), we would have included 65 infants (out of the 68, the 3 incorrect classifications would have been censored). Of those 65 infants, 22 were exposed to an antibiotic. This provides an incidence of antibiotic exposure in the cohort of 22/65 or 33.8%. For any antibiotic exposure for otitis media [not shown], the concordance is still high. In the sensitivity set (n = 68), 4 infants were censored based on unclear antibiotic for otitis media exposure. This left 64 infants that were correctly able to be classified and 19 were exposed to antibiotics for otitis media among them (incidence = 19/64 or 29.7%). Lastly, in the 28-day sensitivity set (n = 68) and censoring for unclear antibiotic exposure for any reason or for unclear otitis media diagnosis, 5 infants would have been censored for antibiotic exposure in general or for antibiotic exposure for otitis media. 20 infants were exposed to antibiotics in general and 19 for otitis media. This makes the incidence 20/63 and 19/63 or 31.7% and 30.2% respectively.

The final NHBCS Antibiotic Exposure Cohort only included infants that were not censored for unclear antibiotic exposure or unclear antibiotic exposure for otitis media ultimately consisting of 183 infants. Of which, 63 were exposed to any antibiotic (incidence = 34.4%), and 58 to an antibiotic for otitis media (incidence = 31.7%).

Thus, our sensitivity analysis showed strong concordance and agreement between the incidence of antibiotic exposures in the sensitivity and ultimate NHBCS Antibiotic Exposure Cohort.

*NHBCS Antibiotic Frequency Cohort:*

Similarly to the sensitivity analysis for the NHBCS Antibiotic Exposure Cohort, we utilized a sensitivity cohort to assess agreement between interview and antibiotic prescriptions data per infant. We included infants in this cohort based on slightly different metrics than the NHBCS Antibiotic Exposure Cohort. Infants were included if they had interview data at 8 and 12 months with the 12-month interview occurring within 28 days of the stool sample along with full pediatric medical record data (n = 64). Additionally, 2 infants were included because they had interview and medical record data beyond the point of their sample collection with no antibiotic exposure. In this sensitivity analysis cohort of 66 infants we found the following results:

|  |  | Pediatric medical record | |
| --- | --- | --- | --- |
|  |  | Antibiotic(s) noted | Antibiotic(s) not noted |
| Interview | Antibiotic(s) noted | 13 | 1 |
|  | Antibiotic(s) not noted | 7 | 45 |

Concordance = (13+45)/66 = 87.8%, Kappa = 68.6%

Note: If multiple antibiotic prescriptions were noted, agreement was only classified if all antibiotic prescriptions were captured in both.

This table demonstrates that there is substantial agreement between methods; however, as expected, some of the antibiotic prescriptions are not captured on the interview. This could be due to a lack of reporting of the antibiotic exposure or that the antibiotic was not given to the infant even when it was prescribed. Given this uncertainty, we decided to include all antibiotic prescriptions. Based on this sensitivity set, if bias was present, we believe this would bias our results towards the null.

**Availability of data and materials**

The whole metagenomic shotgun sequencing samples are available through the National Center for Biotechnology Information (NCBI) Sequence Read Archive: https://www.ncbi.nlm.nih.gov/sra (Accession number: PRJNA296814). Analytic R markdown scripts are available upon request.

**Covariates**

Extensive data on lifestyle, medical history, and environmental exposures were collected from participants in the NHBCS; select covariate data were available for the DIABIMMUNE Study. Across both cohorts, covariates were selected as a result of an *a priori* literature review and a theorized association with antibiotic exposure and the infant gut microbiome. We included gestational age (in weeks)^1^, duration of breastfeeding (in days)^2–4^, delivery mode (vaginal or cesarean)^4–6^, sex (male or female)^7^, infant age at stool sample collection (in days)^5^, and antibiotic usage immediately following birth (yes/no)^8^. Infants were excluded if they had a known antibiotic exposure after the period immediately following birth but before the baseline microbiome sample was collected. Additionally, for NHBCS data, we considered day care attendance by 1 year (yes/no) as a covariate as it had previously been associated with otitis media^9,10^ and we hypothesized that it could have an association with the gut microbiome^11,12^. Adjusted models including infants from both the NHBCS and DIABIMMUNE Study also included a dichotomous covariate to adjust for study origin.

**Stool microbiome assessment**

*NHBCS samples:*

Stool collection and metagenomic DNA shotgun sequencing have been previously completed and described for our cohort^13,14^. In brief, infant stool samples were collected in diapers at approximately 6 weeks and 1 year of age. DNA was extracted from the stool samples with concentrations of DNA samples used for sequencing ranging from 1 ng/ul to 25 ng/ul. Sequencing was conducted at the Marine Biological Laboratory in Woods Hole, MA using established protocols. A Covaris S220 focused ultrasonicator sheared DNA samples to a mean size of 400 base pairs and libraries were constructed using Nugen’s Ovation Ultralow V2 protocol.

All samples were processed using KneadData as part of the bioBakery3 pipeline^15^. This pipeline included samples that were sequenced and merged as paired end reads or samples that were only sequenced as forward reads. Previously, we identified that using both single and paired end reads did not affect associations with early-life factors^13^. Infants were excluded if they did not have both a 6-week and 1-year stool sample. This left 238 infants and 476 total samples. Average sequencing depth for paired (*n* = 237) and single-read (*n* = 239) samples were 61.1 (standard deviation of 21.9) and 24.5 (standard deviation of 10.5) millions of reads respectively.

*DIABIMMUNE Study samples:*

Information regarding stool sample collection and quality control procedures are outlined in the primary research papers by the DIABIMMUNE Study group^16,17^. Stool metagenomic datasets used for this analysis were downloaded directly from the DIABIMMUNE Study website’s Antibiotics Cohort metagenomic forward read sequencing data^18^. Infants were excluded from this analysis if they did not have a 2-month baseline or 1-year sample. Average sequencing depth of the remaining samples (*n* = 66) was 10.6 million reads with a standard deviation of 7.6 million reads.

*Taxonomic and antibiotic resistance gene profiling:*

For samples from both the NHBCS and DIABIMMUNE Studies, taxonomy down to the species level was assigned using MetaPhlAn3^19^ which maps reads from samples to species-specific markers. Only bacterial taxa were considered. Taxa that were identified in less than 1% of samples (assessed in the Antibiotic Exposure and Frequency Cohorts) were excluded in further analyses. After prevalence filtering, the relative abundance at each level of interest (phyla, genera, and species) were normalized to sum to 1.

Markers for ARGs were quantified using ShortBRED^20^. ShortBRED works in two steps. It first creates a set of representative ARG markers from a database of ARG sequences. It then uses this set of highly sensitive and specific markers to probe for sequences in the shotgun metagenomic data that likely confer antibiotic resistance. ARGs of interest were derived from the Comprehensive Antibiotic Resistance Database (CARD)^21^ v3.1.1 antibiotic resistance gene ontologies version dated January 29, 2021 using sequences derived from CARD’s protein FASTA protein homolog model. These proteins of interest were mapped using the shortbred_identify.py script with UniRef90^22^ as the reference protein database^23^. Resulting markers for ARGs were then quantified in the NHBCS sequence datasets using the shortbred_quantify.py script with default settings. Outputs from ShortBRED are normalized for average read length, marker length, and sequencing depth and are represented in reads per kilobase of reference sequence per million samples reads (RPKM). ARGs identified in fewer than 1% of all samples in the Antibiotic Exposure and Frequency Cohorts were removed.

**Statistical analyses**

*Assessing individual off-target microbes and antibiotic resistance genes:*

Our assessment of off-target microbes and ARGs integrated the strengths of the cohort studies; specifically, longitudinal metadata, medical record data, and microbiome data. We were interested in estimating the difference in relative abundance that would have occurred over the follow-up period if an infant had not been exposed to antibiotics.

Estimation of the difference-in-difference estimate was performed using Microbiome Multivariable Associations with Linear Models (MaAsLin2)^24^. Default options were used with the exception of the following, which aided in the interpretation of our results: no transformations were made to assess microbial abundance calculations and no transformations or normalizations were used for ARG assessment. A 95% confidence interval was computed from the output for the difference-in-difference effect estimate’s standard error in MaAsLin2. Our basic formula was:

*Abundance ~ (intercept) + sample age (in days) + antibiotic exposure + antibiotic exposure* $\times$ *sampling interval (baseline vs 1 year) + covariates + (1 | infant)*

Where *abundance* is the relative abundance of the microbe or ARG in each sample, *sample age* is the time in days of the sample collection, *antibiotic exposure* is a dichotomous indicator of if the infant was exposed or not exposed to an antibiotic ever, and *sampling interval* is a binary variable used to denote that the measurement was from baseline (~ 6-weeks or 2 months) or 1-year. *Antibiotic exposure* $\times$ *sampling interval* represents the difference-in-difference estimate or interaction between exposure and time interval. We considered intercept random effects at the infant level *(1| infant)*.

*Modeling antibiotic exposure on overall antibiotic resistance gene abundance:*

To estimate difference-in-difference estimates for the overall ARG abundance per sample, we ran fixed-effects linear regression models with ARG abundance in RPKM as the outcome.

*Overall ARG abundance ~ (intercept) + sample age (in days) + antibiotic exposure + antibiotic exposure* $\times$ *sampling interval (baseline vs 1 year) + covariates*

The same covariates as above were included. Difference-in-difference estimates with 95% confidence intervals are reported.

*Species and antibiotic resistance gene correlation analysis*

Additional analyses examined associations between off-target microbes and ARGs; we used HAllA version 0.8.18 to identify blocks of species and ARGs that were associated with each other^25^. HAllA uses a three-step system that calculates pairwise associations between ‘omics datasets, hierarchically clusters each dataset, and then iteratively identifies associated blocks of features. In particular, we were interested in assessing correlation based on the difference between the baseline and 1-year relative abundance of each species and ARG. Species and ARGs with greater than a 0.5% mean abundance in the Antibiotic Exposure Cohort were included. Species and ARG abundances were centered log-ratio transformed using the microbiome^26^ package to account for relative abundance data. Samples were stratified by antibiotic exposure status by 1 year before running them through HAllA. Spearman correlation with a Benjamini-Hochberg correction of 0.05 was used to associate features and evaluate statistical significance.

*Within-sample (alpha) diversity assessment:*

To estimate the richness and evenness of samples, Shannon alpha diversity at the species level was computed for each sample. Similar to relative abundance assessments, we were interested in estimating the difference in the change in Shannon index that would have occurred over the follow-up period if an infant had not been exposed to antibiotics. Using a linear mixed-effects model implemented with the *lmer()* function in the R package lme4^27^, we evaluated the difference-in-difference estimate for alpha diversity according to the following formula:

*Shannon index ~ (intercept) + sample age (in days) + antibiotic exposure + antibiotic exposure* $\times$ *sampling interval (baseline vs 1 year) + covariates + (1 | infant)*

Where *Shannon index* is the alpha diversity of each sample and the remaining variables are the same as the off-target microbe and ARG abundance calculation.

*Between sample (beta) diversity assessment:*

We analyzed the between sample or beta diversity of bacterial species in infant stool samples in the study. Considering the compositional nature of the data, the relative abundance of species in each sample was centered log-ratio transformed and the distance between samples was measured using the Euclidean distance. This approach, akin to the Aitchison distance, can outperform common metrics such as the Bray-Curtis dissimilarity metric for compositional data and is a true linear difference^28,29^. Differences between samples were visualized using the first 2 components of the principal components analysis. Similar to ^30^, we were interested in how microbiome composition varied by the interaction between an exposure and age. Specifically, we were interested in understanding how compositional variation with respect to antibiotic exposure is affected by compositional variation by age. Dispersion of samples by covariates were analyzed for statistical significance using PERMANOVA models with the *adonis2()* function in the vegan^31^ package. 1000 permutations were run for each model. Samples were removed prior to running the PERMANOVA if they had a missing value for breastfeeding duration.

*Model sensitivity analyses:*

In addition to crude and adjusted models for abundance and diversity metrics, we estimated difference-in-difference or interaction estimates for two other antibiotic exposure scenarios. One set of sensitivity models considered antibiotic exposure specifically for otitis media with an added variable to account for antibiotic exposures not for otitis media (tested with the Antibiotic Exposure Cohort). In the other model sets, infants were only considered exposed if they received two or more antibiotic prescriptions (tested with the Antibiotic Frequency Cohort).

Based on assumptions of the difference-in-difference modeling approach, we stratified infants in the NHBCS cohort based on day care attendance. Difference-in-difference regression was used to evaluate if antibiotic exposure impacted abundance or diversity metrics differentially by the two groups.

All statistical analyses were performed using R version 3.6.0^32^.

**Difference-in-differences assumptions and sensitivity analyses**

*Difference-in-differences assumptions:*

In order for the difference-in-differences approach to represent an unbiased amount of change in the population due to an antibiotic, a few assumptions must be met^33,34^.

The first assumption (often called the strict exogeneity assumption) is that the intervention cannot be associated with the outcome at baseline. While this may *not* be true if we were considering microbes targeted for treatment by the antibiotic exposures, in this case we were only interested in off-target microbes and antibiotic resistance genes.

A second assumption (often called the positivity assumption) is that all people in the study could have been given the intervention. We assume in this study that, since all infants are involved in the medical system to participate in cohort studies, that they all have access to antibiotics.

The last assumption is the parallel trends assumption. This assumption says that there are no unmeasured variables that are both time and group varying. Graphically this can be depicted using two lines to represent the relative abundance of a microbe or ARG among those exposed and unexposed prior to the intervention. If the lines are not parallel, the assumption is not met. These lines are often depicted as linear, but this is not a prerequisite. While this assumption is challenging to address with data at only 2 time points, care has been taken to evaluate variables that could be considered time and group variant. Unless otherwise mentioned, we assume that covariates are time invariant and that the effect of the covariates on our outcome at baseline is also time invariant. This is why we did not assess the interaction between time and other covariates.

*Sensitivity analyses:*

In particular, we explored sample age, duration of breastfeeding, and exposure to day care by 1 year due to time variance (Supplemental tables included below). Sample age and duration of breastfeeding did not vary significantly by group in the Antibiotic Exposure Cohort (n = 216; Kruskal-Wallis *p*-value in baseline and one year > 0.05). However, at baseline, infants eventually exposed to an antibiotic were breastfed slightly longer than infants not exposed to an antibiotic (48.6 days vs. 44.8 days, Kruskal-Wallis p-value = 0.054). Due to this small difference in days breastfeeding, we did not consider this to be a major factor affecting results and did not stratify analyses based on duration of breastfeeding. Since we only had exposure to daycare available in the NHBCS cohorts, we only explored this in NHBCS infants (Supplemental table included below). Among the 183 infants in the Antibiotic Exposure Cohort, infants that were given an antibiotic were also more likely to attend daycare. Thus, in order to tease apart the effects of daycare and antibiotic exposure, we conducted sensitivity analyses among infants that attended and didn’t attend daycare by 1 year.

Exposure to antibiotic by 1 year in Antibiotic Exposure Cohort

|  | **Not exposed** | **Exposed** | **Overall** |
| --- | --- | --- | --- |
|  | **N = 140** | **N = 76** | **N = 216** |
| **Sample age in days of baseline sample** |  |  |  |
| Mean (SD) | 48.1 (15.6) | 49.8 (17.4) | 48.7 (16.2) |
| Median [Min, Max] | 44.0 [28.0, 150] | 45.0 [17.0, 150] | 44.0 [17.0, 150] |
| **Sample age in days of one year sample** |  |  |  |
| Mean (SD) | 368 (27.1) | 375 (28.3) | 371 (27.6) |
| Median [Min, Max] | 368 [319, 570] | 371 [325, 477] | 369 [319, 570] |
| **Duration of breastfeeding in days at baseline** |  |  |  |
| Mean (SD) | 44.8 (19.0) | 48.6 (18.6) | 46.2 (18.9) |
| Median [Min, Max] | 43.0 [0, 150] | 45.0 [0, 150] | 44.0 [0, 150] |
| Missing | 1 (0.7%) | 0 (0%) | 1 (0.5%) |
| **Duration of breastfeeding in days at 1 year** |  |  |  |
| Mean (SD) | 245 (134) | 265 (117) | 252 (128) |
| Median [Min, Max] | 317 [0, 449] | 304 [0, 392] | 306 [0, 449] |
| **Delivery mode** |  |  |  |
| Vaginal | 101 (72.1%) | 59 (77.6%) | 160 (74.1%) |
| C-section | 39 (27.9%) | 17 (22.4%) | 56 (25.9%) |
| **Sex** |  |  |  |
| Female | 55 (39.3%) | 29 (38.2%) | 84 (38.9%) |
| Male | 85 (60.7%) | 47 (61.8%) | 132 (61.1%) |
| **Gestational age (in weeks)** |  |  |  |
| Mean (SD) | 39.1 (1.66) | 39.0 (1.72) | 39.1 (1.68) |
| Median [Min, Max] | 39.1 [29.1, 42.6] | 39.1 [33.3, 41.9] | 39.1 [29.1, 42.6] |

Descriptive overview of infants in New Hampshire Birth Cohort Study Antibiotic Exposure Cohort

|  | **Unexposed to antibiotic** | **Exposed to antibiotic** | **Overall** |
| --- | --- | --- | --- |
|  | **(N=120)** | **(N=63)** | **(N=183)** |
| **Sample age at 6-week sample (days)** |  |  |  |
| Mean (SD) | 45.7 (14.7) | 46.2 (15.1) | 45.8 (14.8) |
| Median [Min, Max] | 43.0 [28.0, 150] | 44.0 [17.0, 150] | 43.0 [17.0, 150] |
| **Sample age of 1 year samples (days)** |  |  |  |
| Mean (SD) | 369 (28.5) | 376 (30.2) | 371 (29.2) |
| Median [Min, Max] | 369 [320, 570] | 371 [325, 477] | 370 [320, 570] |
| **Maternal BMI (kg/m2)** |  |  |  |
| Mean (SD) | 26.1 (5.77) | 26.5 (5.97) | 26.2 (5.83) |
| Median [Min, Max] | 24.4 [17.4, 48.2] | 24.9 [19.0, 47.2] | 24.8 [17.4, 48.2] |
| **Delivery method** |  |  |  |
| Vaginal delivery | 84 (70.0%) | 47 (74.6%) | 131 (71.6%) |
| C-section | 36 (30.0%) | 16 (25.4%) | 52 (28.4%) |
| **Gestational age at birth (weeks)** |  |  |  |
| Mean (SD) | 39.0 (1.66) | 38.9 (1.72) | 38.9 (1.67) |
| Median [Min, Max] | 39.0 [29.1, 41.1] | 39.0 [33.3, 41.9] | 39.0 [29.1, 41.9] |
| **Parity** |  |  |  |
| 0 | 61 (50.8%) | 28 (44.4%) | 89 (48.6%) |
| 1+ | 54 (45.0%) | 34 (54.0%) | 88 (48.1%) |
| Missing | 5 (4.2%) | 1 (1.6%) | 6 (3.3%) |
| **Attend day care by 1 year** |  |  |  |
| No | 68 (56.7%) | 23 (36.5%) | 91 (49.7%) |
| Yes | 40 (33.3%) | 36 (57.1%) | 76 (41.5%) |
| Missing | 12 (10.0%) | 4 (6.3%) | 16 (8.7%) |
| **Breastfeeding duration at 6 weeks (days)** |  |  |  |
| Mean (SD) | 41.9 (18.3) | 45.0 (16.6) | 43.0 (17.7) |
| Median [Min, Max] | 42.0 [0, 150] | 43.0 [0, 150] | 43.0 [0, 150] |
| Missing | 1 (0.8%) | 0 (0%) | 1 (0.5%) |
| **Breastfeeding duration at 1 year (days)** |  |  |  |
| Mean (SD) | 238 (139) | 265 (119) | 247 (133) |
| Median [Min, Max] | 312 [0, 449] | 304 [0, 391] | 304 [0, 449] |
| **Sex** |  |  |  |
| Female | 46 (38.3%) | 24 (38.1%) | 70 (38.3%) |
| Male | 74 (61.7%) | 39 (61.9%) | 113 (61.7%) |
| **Maternal education level** |  |  |  |
| High school or equivalent | 11 (9.2%) | 5 (7.9%) | 16 (8.7%) |
| Junior college, technical school, or some college | 20 (16.7%) | 6 (9.5%) | 26 (14.2%) |
| College | 38 (31.7%) | 29 (46.0%) | 67 (36.6%) |
| Post-graduate | 49 (40.8%) | 22 (34.9%) | 71 (38.8%) |
| Missing | 2 (1.7%) | 1 (1.6%) | 3 (1.6%) |
